# Quality of life in adolescents with chronic non-traumatic knee pain: An analysis of 323 adolescents with Patellofemoral Pain and Osgood-Schlatter Disease

**DOI:** 10.1101/2023.01.19.23284544

**Authors:** Chris Djurtoft, Tomer Yona, Ewa Maria Roos, Kristian Thorborg, Per Hölmich, Sten Rasmussen, Jens Lykkegaard Olesen, Michael Skovdal Rathleff.

**Affiliations:** Center for General Practice at Aalborg University, Aalborg, Denmark; Department of Health Science and Technology, Aalborg University, Denmark; Department of Biomedical Engineering, Technion, Israel Institute of Technology, Haifa, Israel; Research Unit for Musculoskeletal Function and Physiotherapy, Center for Muscle and Joint Health, Institute of Sport Science and Clinical Biomechanics, University of Southern Denmark, Odense, Denmark; Sports Orthopedic Research Center – Copenhagen (SORC-C), Hvidovre, Denmark; Physical Medicine and Rehabilitation Research-Copenhagen (PMR-C), Hvidovre, Denmark; Department of Orthopedic Surgery, Copenhagen University Hospital, Hvidovre, Denmark; Department of Clinical Medicine, University of Copenhagen, Copenhagen, Denmark; Department of Clinical Medicine, Aalborg University, Aalborg, Denmark; Department of Orthopedic Surgery, Aalborg University Hospital; Department of Physiotherapy and Occupational Therapy, Aalborg University Hospital, Aalborg, Denmark

**Keywords:** Quality of life, Non-traumatic knee pain, Patellofemoral Pain, Osgood-Schlatter Disease

## Abstract

**Introduction:** We aimed to describe Quality of life (QoL) among adolescents with Patellofemoral Pain (PFP) and Osgood-Schlatter Disease (OSD) according to the QoL subscale of The Knee injury and Osteoarthritis Outcome Score (KOOS) and the EuroQol 5-dimensions (EQ-5D) and to investigate the characteristics associated with QoL.

**Methods:** This individual participant analysis included data from three clinical trials on adolescents diagnosed with PFP or OSD. We relabeled individual data and constructed a single dataset.

**Results:** We included 323 adolescents with PFP or OSD. Total score of KOOS-QoL was 51±18 and total index score for the EQ5D was 0.67±0.21. KOOS-QoL subscale showed that 59% reported being aware of their knee problems daily or constantly, 37% reported severe to extreme lack of confidence in their knees, 27% reported severe to extreme difficulty with their knees, and 20% reported severely or totally modifying their lifestyle to avoid potentially damaging activities to their knee. EQ-5D showed that 77% experienced problems with everyday activities, 47% reported mobility problems, 17% felt worried, sad, or unhappy, and 7% reported problems looking after themselves. Older participants (age 17-19) reported worse QoL than younger participants. None of the other characteristics were associated with QoL.

**Conclusions:** A high proportion of adolescents with chronic non-traumatic knee pain experience low QoL. More than half were aware of their knee problems at least daily, one in three reported a severe lack of confidence in their knee, and one in six felt worried, sad, or unhappy. QoL was lowest among older adolescents.

**Highlights:**

- One in every three adolescents with PFP or OSD reported a severe lack of confidence in their knee.
- Many adolescents reported being sad or worried, and modified their usual activities due to their knee pain.
- Clinicians may extend the scope and include pain management strategies to address psychological perspectives when managing adolescents diagnosed with PFP or OSD.
- Modifiable targets such as adolescents understanding of pain may be a beneficial management strategy to consider in rehabilitation for PFP or OSD.

## Introduction

Chronic musculoskeletal pain (CMP) affects up to one-third of adolescents (age 10-19) ^6,9,34^. During this critical period in life, pain may have detrimental impact on a child’s physical, emotional, and social functioning ^6,8,17^. Non-traumatic knee pain is the most common pain origin among adolescents, with Patellofemoral Pain (PFP) and Osgood-Schlatter Disease (OSD) having the highest prevalence ^34^. PFP and OSD show similar pain patterns, including pain in the anterior part of the knee, and are commonly observed among highly active adolescents ^28,35^. PFP and OSD are often seen as self-limiting, but recent evidence shows that approximately half of the adolescents having knee pain will continue to experience pain after 12 months, and nearly 75% of adolescents will reduce sports participation due to their knee pain ^12,29,32^. The impact of PFP and OSD has significant consequences, as issues extend to basic functional tasks (running and stair-walking), shorter sleeping time, worse sleep quality, and worse quality of life (QoL) than their peers without pain ^12,22,25,29,32^.

A recent individual participants meta-analysis investigating prognostic factors for long-term outcomes demonstrated that 51% of adolescents with non-traumatic knee pain still report knee pain after 12 months ^12^. Notably, low Health-Related Quality of Life (HRQoL) was one of the strongest prognostic factors for poor outcomes and the only identified construct that seems modifiable ^12^. Importantly, modifiable risk factors have high clinical value, as they might help clinicians tailor patient-centered management strategies to enhance rehabilitation success ^23^. However, it is challenging to identify characteristics of predictive value and previous evidence regarding prognostic factors in adolescents CMP is sparse and inconsistent ^12,13,27^.

Despite HRQoL has been suggested as an important modifiable prognostic factor, it has remained unclear which characteristics are associated with this phenomenon in this population ^12,13^. However, QoL is a multidimensional construct that is highly subjective and associated with many aspects in adolescents experiencing CMP, such as stress, loneliness, lower self-efficacy, and lower self-esteem ^16,20,22^. Additionally, identifying ways to manage and improve QoL was recently ranked as the second highest research priority within CMP ^26^, which emphasizes the need for an increased understanding of what influences QoL in adolescents diagnosed with PFP or OSD. This will strengthen our knowledge of this phenomenon and plausibly provide important modifiable targets for current management strategies ^27^.

Combining individual participants’ data from three published original trials on adolescents with PFP or OSD strengthens statistical power and raises an opportunity to explore what influences QoL in a larger group of adolescents with the two most common knee conditions. Therefore, we aimed to 1) describe QoL among adolescents with PFP or OSD according to the EuroQol 5-dimensions (EQ-5D) and The Knee injury and Osteoarthritis Outcome Score (KOOS) QoL and 2) to investigate measures of QoL and its association with characteristics in terms of sports participation, sports activities per week, pain duration, pain intensity, bilateral knee pain, body mass index (BMI), sex, age, and use of pain medication.

## Methods

### Study Design

Our study is reported in accordance with the STROBE statement ^7^. This cross-sectional explorative study was designed as an individual participant analysis of three previous clinical trials on adolescents (defined by the World Health Organization (WHO) as the second decade of life, 10-19 years old ^40^) suffering from chronic non-traumatic knee pain. Trials are registered on clinicaltrials.gov (NCT02799394, NCT02402673, NCT01438762). Signed informed consent was obtained from all participants and all data were stored on a secure fileshare. This study was initiated by the original author group of the original publications ^28,33,36^.

### Description of Included Studies

The three studies recruited adolescents diagnosed with PFP or OSD. Two studies included adolescents with PFP, and one included adolescents with OSD.

### Patient Demographics and Exposures

All studies collected information on sports participation, sports activities per week, pain duration, bilateral knee pain, BMI, sex, age, and use of pain medication; additionally, we collected knee pain intensity (worst pain last week), which was measured using visual analog scale or numeric pain rating scales.

Knee-related QoL was collected by the KOOS questionnaire, which contains five subscales (pain, symptoms, activity in daily living, function in sport and recreation, and knee-related QoL) scored from 0 to 100 (worst to best) ^4,37^. As this analysis’s primary interest was to describe QoL, we collected data from the KOOS-QoL subscale and individual items representing the knee-related QoL. Health-related QoL was measured by the EQ-5D in study one and the EQ-5D youth version in studies two and three. The two versions contain the same items but slightly different wordings to ensure adolescents below the age of 15 also comprehend the language used. The Danish weights were used to calculate the EQ-5D index score (a higher index score equals better QoL) ^49^.

### Data Harmonization and Checking

After acquiring the data, we relabeled individual data sets and recoded them to ensure consistency and construct a single, merged dataset. The individual patient data validity was checked by undertaking completeness and consistency checks on individual participant data to identify missing or invalid (e.g., out of range) items. Missing information or inconsistencies were checked and rectified, as necessary. The original three studies were powered to explore the clinical benefits of a specific management strategy. The sample size was thus fixed by the original studies.

### Data Analysis

Normally distributed continuous variables are described as mean ± standard deviation or 95% confidence intervals. Non-normally distributed data is presented as median and [range]. Categorical variables are expressed as frequencies and percentages.

For Knee-related QoL, we calculated the subscale mean score measured by the KOOS-QoL as recommended where points are summed, converted to percentages of a maximum score and reversed to a 0-100, worst to best scale ^4,37^. In addition, the individual items were divided into scores from Likert scales ^4,37^. All items were answered on a 5-point Likert scale (never/not at all, monthly/mildly, weekly/moderately, daily/severely, constantly/totally/extreme), yielding 0–4 points, with 0 points representing no difficulty ^4,37^. For the QoL measured by the EQ-5D, we present the index score as a general proxy for QoL, the “health profiles”, and the individual five items divided into “no problems” or “any problem”, as suggested in the EQ-5D User Guide ^49^.

Characteristics associated with the KOOS-QoL and EQ-5D are presented using box plots, displaying the range, median, and interquartile range of scores. The horizontal line represents the median score, and each box represents the interquartile range. The “whiskers” signify the lowest and highest score for each group. The circles outside the whiskers represent scores above the 10th and 90th percentiles, respectively; these scores fall outside the regular range of values in a distribution and are regarded as outliers. For pragmatic reasons, we divided BMI into five groups (1: <18.5 - 2: 18.5 to 24.9 – 3: 25 to 29.9 – 4: 30 to 39.9 – and 5: ≥40), according to the description provided by the WHO ^5^. No participants had a BMI >40, therefore this category is not represented. We divided knee pain duration into three groups (1: 0-6 month – 2: 6-12 month – and 3: ≥12 month), as recommended ^1^, and lastly, knee pain-intensity was divided into quartiles (Q1: 0-.25 – Q2: 0.25-0.5 – Q3: 0.5-0.75 and Q4: 0,75-1). All analyses were carried out using Statistical Package for the Social Sciences (SPSS), version 25.

## Results

### Characteristics of participants

A total of 323 participants were included in the dataset; 237 (73%) were girls, 241 (75%) reported bilateral knee pain, and their ages ranged from 10 to 19 (Table 1). Of all participants, 243 (75%) engaged in sports activities at least twice a week, and 66 (21%) used pain medication due to knee pain.

**Table 1.** Characteristics of included participants and studies. Descriptive data are mean (SD) unless otherwise stated.

|  | Study 1<br>Patellofemoral<br>pain (n=121) | Study 2<br>Osgood<br>Schlatter (n=51) | Study 3<br>Patellofemoral<br>Pain (n=151) | All<br>participants<br>(n=323) |
| --- | --- | --- | --- | --- |
| Age (years) | 17 [15-19] | 13 [10-15] | 13 [10-14] | 14 [10-19] |
| Female sex, n (%) | 97 (80.2%) | 25 (49%) | 115 (76.2%) | 237 (73.4%) |
| Body mass index, (kg/m<sup>2</sup>) | 21.6 [17.1-32.8] | 20.3 [14.7-27.6] | 19 [14-28] | 19.8 [14-32.8] |
| Pain duration (months) | 39 [2-192] | 18 [2-60] | 18 [2-78] | 24 [2-192] |
| Worst pain last week (VAS/NRS) | 48 [3-97] | 70 [0-100] | 60 [0-100] | 55 [0-100] |
| Bilateral knee pain, n (%) | 95 (78.5) | 35 (68.6) | 111 (73.5) | 241 (74.8) |
| Use of pain medication, n (%) | 24 (19.8) | 6 (11.8) | 36 (23.8) | 66 (20.6) |
| Sport activities (yes), n (%) | 81 (66.9) | 38 (74.5) | 124 (82.1) | 243 (75.2) |
| Sport activities per week | 2 [0-7] | 2 [0-6] | 2 [0-7] | 2 [0-7] |

Continuous variables are presented as median and [range]. Nominal variables are presented as number (percentages). Abbreviations; SD, Standard deviation; NRS, numeric rating scale; VAS, visual analogue scale.

### KOOS-QoL subscale scores

KOOS-QoL subscale score for the 323 participants was 51±18. The most prominent item of KOOS-QoL was awareness of the knee problem, with 191 (59%) reporting daily or constant awareness (Table 2). Adolescents that reported severe to extreme lack of confidence in their knees were 119 (36%) and 87 (27%) reported severe to extreme difficulty with their knees. Lastly, 64 (20%) reported severely or totally modifying their lifestyle to avoid potentially damaging activities.

**TABLE 2.** Participants Knee injury and Osteoarthritis Outcome Score (KOOS) QoL subscale

|  | **Study 1 (n=121)**<br>**Patellofemoral pain** | **Study 2 (n=51)**<br>**Osgood Schlatter** | **Study 3 (n=144)**<br>**Patellofemoral Pain** | **All participants**<br>**(n=323)** |
| --- | --- | --- | --- | --- |
| **Q1. How often are you aware of your knee problem?** |  |  |  |  |
| Never | 2 (1.7) | 1 (2) | 0 | 3 (0.9) |
| Monthly | 7 (5.8) | 1 (2) | 3 (2) | 11 (3.4) |
| Weekly | 62 (51.2) | 9 (17.6) | 40 (26.5) | 111 (34.4) |
| Daily | 47 (38.8) | 32 (62.7) | 87 (57.6) | 166 (51.4) |
| Constantly | 3 (2.5) | 8 (15.7) | 14 (9.3) | 25 (7.7) |
| Missing | 0 | 0 | 7 (4.6) | 7 (2.2) |
| **Q2. Have you modified your lifestyle to avoid potentially damaging activities to your knee?** |  |  |  |  |
| Not at all | 40 (33.1) | 7 (13.7) | 36 (26.8) | 83 (25.7) |
| Mildly | 53 (43.8) | 15 (29.4) | 54 (35.8) | 122 (37.8) |
| Moderately | 9 (7.4) | 14 (27.5) | 24 (15.9) | 47 (14.6) |
| Severely | 15 (12.4) | 10 (19.6) | 25 (16.6) | 50 (15.5) |
| Totally | 4 (3.3) | 5 (9.8) | 5 (3.3) | 14 (4.3) |
| Missing | 0 | 0 | 7 (4.6) | 7 (2.2) |
| **Q3. How much are you troubled with lack of confidence in your knee?** |  |  |  |  |
| Not at all | 7 (5.8) | 4 (7.8) | 27 (17.9) | 38 (11.8) |
| Mildly | 33 (27.3) | 12 (23.5) | 28 (18.5) | 73 (22.6) |
| Moderately | 35 (28.9) | 12 (23.5) | 39 (25.8) | 86 (26.6) |
| Severely | 43 (35.3) | 21 (41.2) | 41 (27.2) | 105 (32.5) |
| Extremely | 3 (2.5) | 2 (3.9) | 9 (6) | 14 (4.3) |
| Missing | 0 | 0 | 7 (4.6) | 7 (2.2) |
| **Q4. In general, how much difficulty do you have with your knee?** |  |  |  |  |
| None | 4 (3.3) | 4 (7.8) | 10 (6.6) | 18 (5.6) |
| Mild | 36 (29.8) | 5 (9.8) | 21 (13.9) | 62 (19.2) |
| Moderate | 62 (51.2) | 17 (33.3) | 70 (46.4) | 149 (46.1) |
| Severe | 18 (14.9) | 24 (47.1) | 39 (25.8) | 81 (25.1) |
| Extreme | 1 (0.8) | 1 (2) | 4 (2.6) | 6 (1.9) |
| Missing | 0 | 0 | 7 (4.6) | 7 (2.2) |
| **KOOS QoL subscale Mean Score (Mean, SD)** | **55±16** | **43±16** | **51±19** | **51.4±18** |

Abbreviations; KOOS, Knee injury and Osteoarthritis Outcome Score; SD, Standard deviation; QoL, Quality of life.

### EQ-5D Scores

The total index score of the EQ5D was 0.67± 0.21. Pain or discomfort was the most affected item on EQ-5D (Table 3), with 297 (92%) reporting problems. A total of 251 (77%) participants reported problems with usual activities and 153 (47%) had problems with mobility. Feeling worried, sad, or unhappy was noted in 55 (17%) participants, and 23 (7%) reported having problems looking after themselves.

**TABLE 3.** EQ-5D data, showing frequencies and proportions by dimension and severity level.

|  | Study 1 (n=121) | Study 2 (n=51) | Study 3 (n=151) | All participants (n=323) |
| --- | --- | --- | --- | --- |
| **Mobility** |  |  |  |  |
| No problems | 70 (57.9) | 25 (49) | 68 (45) | 163 (50.5) |
| Some problems | 51 (42.1) | 26 (51) | 68 (45) | 145 (44.9) |
| A lot of problems | 0 | 0 | 8 (5.3) | 8 (2.5) |
| Missing | 0 | 0 | 7 (4.6) | 7 (2.2) |
| **Looking After Myself** |  |  |  |  |
| No problems | 118 (97.5) | 45 (88.2) | 130 (86.1) | 293 (90.7) |
| Some problems | 3 (2.5) | 6 (11.8) | 14 (.93) | 23 (7.1) |
| A lot of problems | 0 | 0 | 0 | 0 |
| Missing | 0 | 0 | 7 (4.6) | 7 (2.2) |
| **Doing Usual Activities** |  |  |  |  |
| No problems | 28 (23.1) | 7 (13.7) | 31 (20.5) | 66 (20.4) |
| Some problems | 90 (74.4) | 36 (70.6) | 91 (60.3) | 217 (67.2) |
| A lot of problems | 3 (2.5) | 8 (15.7) | 22 (14.6) | 33 (10.2) |
| Missing | 0 | 0 | 7 (4.6) | 7 (2.2) |
| **Having Pain or Discomfort** |  |  |  |  |
| No problems | 9 (7.4) | 1 (2) | 9 (6) | 19 (5.9) |
| Some problems | 107 (88.4) | 38 (74.5) | 98 (64.9) | 243 (75.2) |
| A lot of problems | 5 (4.1) | 12 (23.5) | 37 (24.5) | 54 (16.7) |
| Missing | 0 | 0 | 7 (4.6) | 7 (2.2) |
| **Feeling Worried, Sad or Unhappy** |  |  |  |  |
| No problems | 106 (87.6) | 40 (78.4) | 115 (76.2) | 261 (80.8) |
| Some problems | 14 (11.6) | 9 (17.6) | 26 (17.2) | 49 (15.2) |
| A lot of problems | 1 (0.8) | 2 (3.9) | 3 (2) | 6 (1.9) |
| Missing | 0 | 0 | 7 (4.6) | 7 (2.2) |
| **EQ5D Index-score (Mean, SD)** | 0.74 (0.12) | 0.63 (0.23) | 0.62 (0.25) | 0.67 (0.21) |

Abbreviations; EQ5D, EuroQol 5-dimensions; SD, Standard deviation.

### Characteristics associated with QoL

Based on visual interpretation, we examined the association between measures of QoL and included variables. Visual inspection showed no linear association between weekly sports participation and KOOS-QoL or EQ-5D scores (Figure 1). Adolescents participating in little or no sport and those participating six times weekly or more reported the lowest QoL. In addition, adolescents reported being sports active had equal level of QoL as those who did not participate in sports. Visual inspection revealed no association between knee pain intensity and KOOS-QoL or EQ-5D scores. For pain duration, no association was observed at any time-points. Adolescents reporting use of pain medication did not score lower quality of life compared to adolescents reporting no use of pain medication.

**FIGURE 1.**
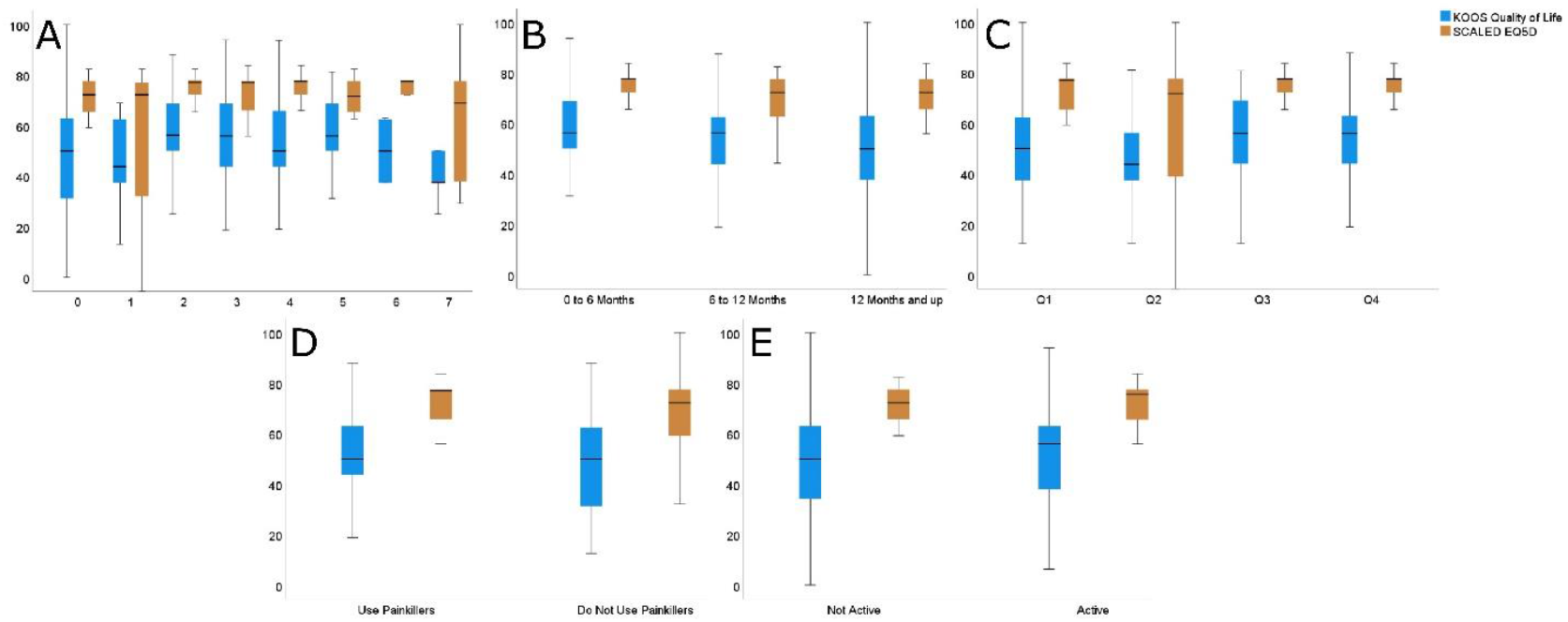
EQ-5D-Y index and KOOS QoL scores by sport participation per week. The error bars represent the 95% confidence intervals. Abbreviations; EQ5D, EuroQol 5-dimensions; KOOS, Knee injury and Osteoarthritis Outcome Score; Q, Quartiles; QoL, Quality of life. A= Sports activities per week; B= Knee pain duration; C= Knee pain intensity; D= Pain medication; E= Sports participation.

Sex was not associated with lower QoL, and older participants (age 17-19) reported lower QoL than younger participants (Figure 2). Adolescents having bilateral knee pain reported similar levels of QoL as adolescents having unilateral knee pain, and lastly, for BMI, we observed no difference in perceived QoL between any of the predefined groups.

**FIGURE 2.**
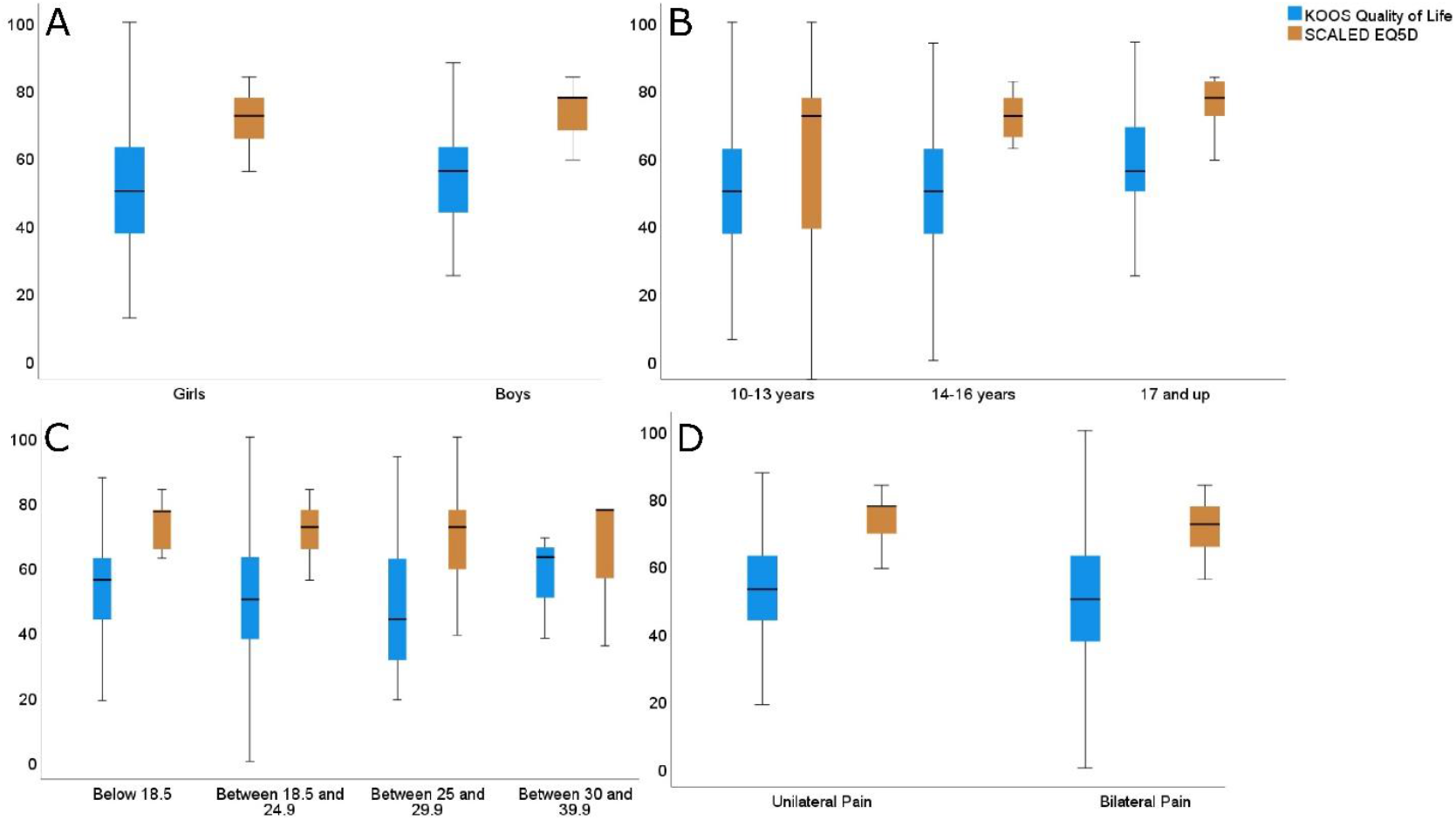
EQ-5D-Y index and KOOS QoL scores by gender. The error bars represent the 95% confidence intervals. Abbreviations; EQ5D, EuroQol 5-dimensions; KOOS, Knee injury and Osteoarthritis Outcome Score; QoL, Quality of life; A= Sex; B= Age; C= Body mass index (BMI); D= Bilateral knee pain.

## Discussion

We found that many adolescents reported a lack of confidence in their knee, often felt sad or worried, and modified their usual activities due to their knee pain. Our findings contribute to the growing evidence base that PFP and OSD are associated with negative psychological impact. This may contribute to the complex experience of CMP ^17,42^. These findings extend our current knowledge and highlight the negative impact chronic non-traumatic knee pain can have on adolescents. This may prompt clinicians with new targets during rehabilitation, beyond the functional deficits these young individuals diagnosed with PFP or OSD may experience.

### Explanation of Findings

Of 323 adolescents, 77% changed their usual activities to avoid what they perceived as damaging to their knees, and 36% reported severe to extreme lack of confidence in their knees. Our findings are consistent with a previous study among adolescents diagnosed with either PFP or OSD which showed more than 50% reported their reduced sports participation, due to *“pain”*, and “*I am afraid to damage my knee”* ^35^. Adolescents understanding of their pain condition may have a direct impact on how they interpret and manage their pain ^8,17,42^. Some adolescents with a lived experience of CMP ascribe positive changes in understanding of pain (i.e., pain as non-threatening) as important contributors towards their recovery ^8,15,17^. Johansen et al. recently showed that the individual perceptions of ‘why do I have pain’ (i.e., functional theories) are strong drivers of behavior in young adults with non-traumatic knee pain underlining that knowledge about their condition is important ^14^. Adolescents with more knowledge about causes of their pain, also showed increased ability to accept, cope and control their knee pain ^14^. This helped them to adjust their perceived QoL and may explain why QoL is associated with prognosis ^12,14,17^. Combined with our findings, it might suggest that these psychological aspects, such as negative beliefs about pain play an important role in rehabilitation for adolescents.

Our findings raise important questions about current rehabilitation for adolescents experiencing chronic non-traumatic knee pain. Traditionally, PFP and OSD management has focused on physical interventions, such as exercise, resistance training, taping, and stretching ^3,41,47,48^. However, it is unclear if these interventions target the QoL dimensions we have identified ^6,24,48^. Previous research on adolescents with non-traumatic knee pain show that over the course of treatment QoL does not improve to the same extent as pain and function ^28,30,31,33^. This opens up the question if we need to reconsider content and focus during rehabilitation of adolescents with PFP or OSD ^2,6^. Our findings suggest evidence-based management for adolescents with PFP and OSD may benefit from an even broader range of modifiable targets, such as understanding of pain, low confidence, negative emotions, and fear of damaging the knees to improve QoL and potentially their prognosis. Nonetheless, we cannot exclude any possibilities that other relevant contributing dimensions may play a significant role during rehabilitation since these were not measured. The cross-sectional design of this study prevents us from both causal conclusions and the longitudinal understanding; we urge future studies to examine this further.

In the current study, we observed that over half (59%) of adolescents with PFP or OSD reported daily or constant awareness of their knee pain and almost one fifth felt worried, sad, or unhappy. However, these negative beliefs and important dimensions of QoL were not associated with duration of knee pain, although this being a well-known determinant for prognosis ^12^. Measures of QoL have been suggested as a proxy for current state in life in relation to self-evaluated health and therefore, prone to significant change in meaning over time ^39,45^. Adolescents’ preferences and goals change over the course of time and are highly dependent of the perceived stage of rehabilitation ^14,15,46^. Sprangers and Schwartz describe this phenomenon as *response shift*; a result of changing personal norms, values, or conceptualizing on QoL throughout the course of a condition ^43,45^. The response shift illustrates individual capacity to adapt to their context (i.e., acceptance of pain), shifting focus away from one cannot (for now), to what they can do to exert control of pain in their current situation ^43,45^. This aligns with qualitative evidence where some adolescents with CMP ascribe that enhanced knowledge of pain legitimize self-management behaviors that makes it easier to accept limitations, justify actions to other peers, prioritize new life goals and thereby accomplish satisfactory level of perceived QoL ^8,14,15,38^. The temporal and adaptive nature of this phenomenon may explain why perceived QoL differs from other outcomes in the long-term ^45^.

We did not observe any strong association between measures of QoL and characteristics, only older adolescents (≥17 years) reported lower QoL compared with younger adolescents. However, a common theme across our results were the large data variability, which emphasizes the heterogeneity of self-reported QoL in this population. This is consistent with previous research on traumatic knee injuries in young individuals ^44^. They showed that QoL was very variable and that several individual psychological, social, and contextual dimensions related to QoL influenced and changed differently across all staged during rehabilitation ^44^. These issues reflect the diverse impact and needs during rehabilitation these young individuals may experience. Adolescents’ diverse clinical presentation may provide a plausible explanation for current insufficient evidence of recommending specific treatments in the context of non-traumatic knee pain ^24,48^. It has been suggested that current management for adolescents CMP may benefit from higher degree of personalization that fit into each individual context ^6,21^. The current study adds new insights into this field of adolescents suffering from PFP or OSD. Our results may help tackle some of the challenges clinicians face when tailoring rehabilitation to fit individual adolescent.

### Clinical Implications

Our findings may help increase clinicians’ awareness of psychological perspectives in adolescents diagnosed with PFP or OSD. It is our clear recommendation that clinicians keep our results in mind when providing personalized evidence-based pain assessment and subsequent management for adolescents with PFP or OSD. There may be unique management options to consider in clinical settings. Future management might benefit from clinicians addressing the whole biopsychosocial perspective tailored to the individual’s context ^6,24,48^. Lancet Child & Adolescent Health Commission emphasized the importance of intervening earlier to ensure young people get the proper management at the right time. Early identification of these modifiable targets may help clinicians tailor personalized rehabilitation to optimize relevant outcomes, such as adolescents perceived QoL ^6^.

### Strength and limitations

Strengths of our study include the large sample, the broad age range of participants, and the use of two complementary measures of QoL; one knee-specific and one generic measure which yielded similar results ^11^. The following limitations should also be considered in interpreting the findings of our study. First, our results rely on a cross-sectional explorative study design. Therefore, our results should be seen as hypotheses generating rather than proving any causal or temporal relationships between measures of QoL and characteristics. Second, recent studies emphasize that current measures of QoL are prone to conceptual and methodological challenges and may lack validity of the target construct ^10,11,19^. This means our data must be interpreted cautiously as we may have missed important dimensions of QoL ^11,18,19,45^.

### Future Research

Using qualitative methods, future studies should explore the questions of QoL to capture the whole experience of chronic non-traumatic knee pain and understand how and why changes in perception occur over time. We encourage future studies to use a longitudinal design to understand how QoL can be modified and explore co-occurring dimensions associated with QoL as well as changes in QoL. This will provide essential knowledge to further understand the complexity of living well with (or despite) the knee pain and potentially develop new management strategies for these individuals.

### Conclusion

A high proportion of adolescents with chronic non-traumatic knee pain experience low QoL. More than half were aware of their knee problems at least daily, one in three reported a severe lack of confidence in their knee, and one in six felt worried, sad, or unhappy. Boys reported lower QoL than girls and QoL were lowest among older adolescents. These results may inform relevant targets in rehabilitation for adolescent non-traumatic knee pain.

## Data Availability

Data are available upon request to corresponding author.

## Acknowledgements, financial disclosure, and conflict of interest

The funders had no role in study design, data collection and analysis, decision to publish, or preparation of the manuscript. All authors certify that they have no affiliations with or involvement in any organization or entity with any financial interest or nonfinancial interest in the subject matter or materials discussed in this manuscript.

## Notes

### Competing Interest Statement

The authors have declared no competing interest.

### Funding Statement

This study was funded by the Danish Independent Research Foundation (grant DFF-4004-00247) and the Danish Physiotherapists Research Foundation. The authors certify that they have no affiliations with or financial involvement in any organization or entity with a direct financial interest in the subject matter or materials discussed in the article.

### Author Declarations

Ethics Committee of North Denmark Region. This study was exempt from a full ethical approval by The North Denmark Region Committee on Health Research Ethics.

