## Supplementary figures and images for "Quality of life in adolescents with chronic non-traumatic knee pain: An analysis of 323 adolescents with Patellofemoral Pain and Osgood-Schlatter Disease"

### Figure 1

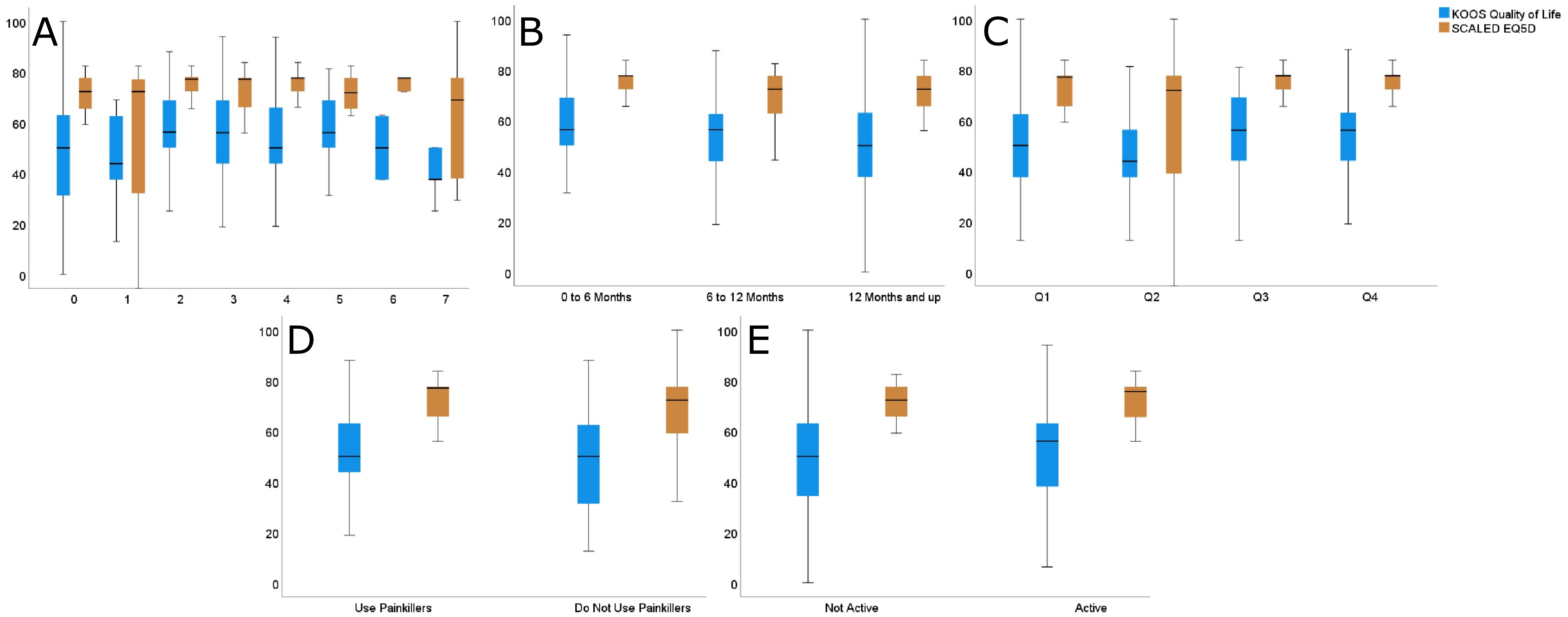

### Figure 2

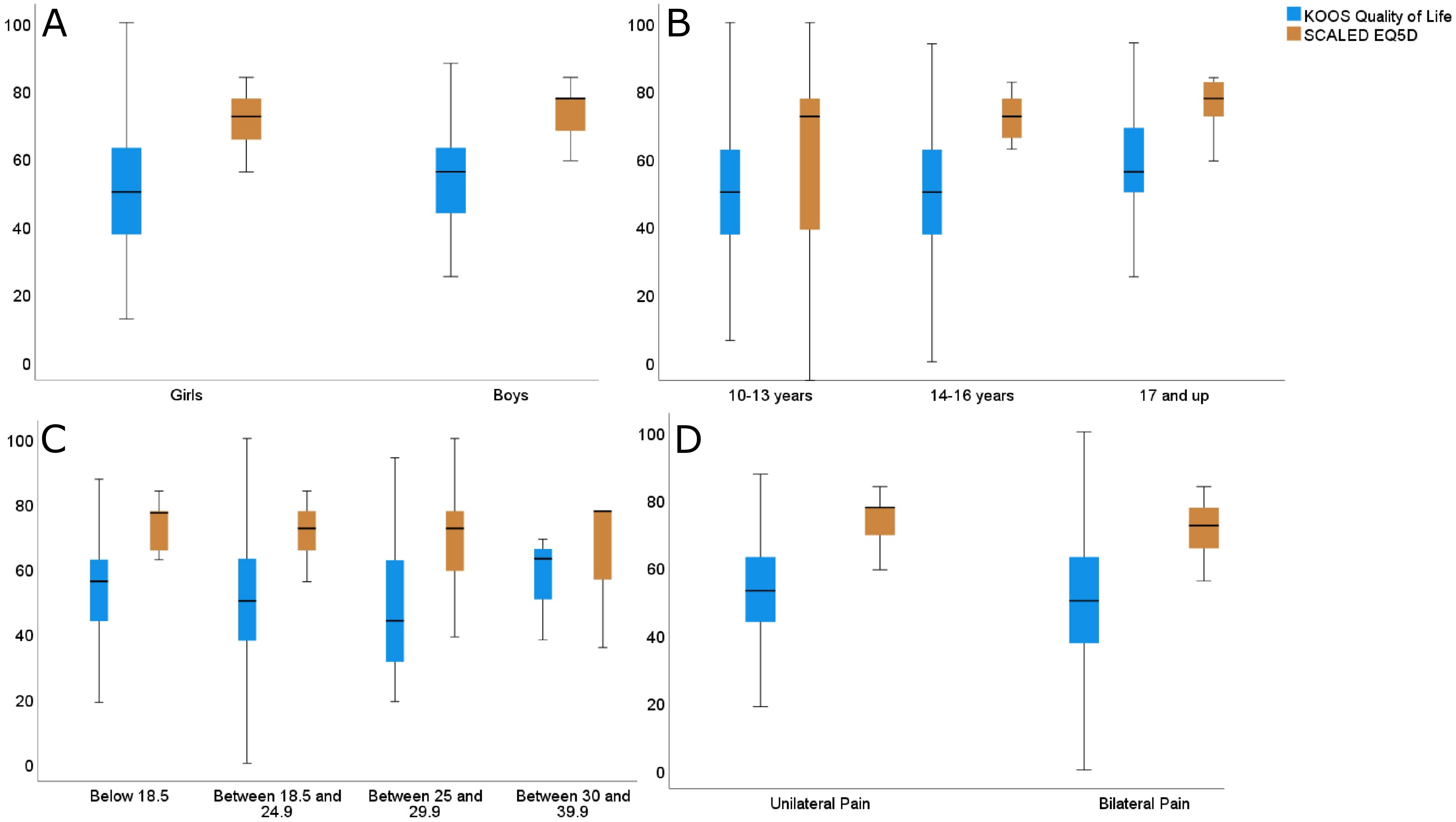
